# Workplace risk management for SARS-CoV-2: a three-step early intervention strategy for effective containment of infection chains with special regards to virus variants with increased infectivity

**DOI:** 10.1101/2021.07.21.21260756

**Authors:** Andreas Paaßen, Laura Anderle, Karsten John, Sebastian Wilbrand

## Abstract

**Background:** Priority during the SARS-CoV2 pandemic is that employees need to be protected from infection risks and business activities need to be ensured. New virus variants with increased infection risks require an evolved risk strategy.

**Material and methods:** Several standard measures such as testing, isolation and quarantine are combined to a novel risk strategy. Epidemiological model calculations and scientific knowledge about the course of SARS-CoV2 infectivity are used to optimize this strategy. The procedure is implemented in an easy-to-use calculator based on Excel.

**Layout in practice and results:** Alternative combinations of measures and practical aspects are discussed. Example calculations are used to demonstrate the effect of the discussed measures.

**Conclusion:** That quarantine calculator derived from these principles enables even non-specialists to perform a differentiated risk analysis and to introduce optimized measures. Targeted testing routines and alternative measures ensure staff availability.

## Introduction

In a workplace setting, the priority during the SARS-CoV2 pandemic is to protect employees and nonemployees on the one hand and to ensure business activity on the other hand. High numbers of new infections overload company structures and resources and continuously introduce contagion risks into facilities. In this context, an internal chain of infection is the greatest threat to the continuation of business activities, as it can affect many employees simultaneously in a short period of time. With their ability to cause eruptive outbreaks, the new virus variants require an evolved risk strategy.

The two most important aspects of addressing this are (1) minimizing infection risks by effective protective measures and (2) an efficient isolation and quarantine management using contact tracing. Therein, companies have effective intervention options at their disposal, such as the binding directive of protective measures and the possibility of renouncing to on-site work and thus removing in-house infection risks. Using these intervention possibilities and an evolved risk management can help operations keep intra-company infection risks for employees very low, as compared to external risks.

In parallel to reliably preventing infection chains, companies have interest in ensuring a high level of operational staff availability to continue business operations. With special regards to highly infective virus variants, there is a strong demand for the implementation of advanced and alternative measures in isolation and quarantine management.

The isolation and risk management strategy discussed in the following relies on epidemiologic models that allow for detailed risk assessment. It has been developed in parallel to its assessment in practice at Marl Chemical Park, a large German chemical site with approximately 10 000 employees and around 3000 external employees working in investment projects.

Very short response times and timely and precisely targeted measures are decisive factors of success for the presented procedures. A calculation tool based on Excel and epidemiologic models developed for this aim is presented and discussed in this paper.

Juridical aspects of measures such as isolation of infective persons and quarantine for contact persons will not be discussed in the following. Depending on regional or state regulations, these measures may be obligatory by law or by administrative directives. Moreover, employers can release their employee from work duties or assign mobile work. Self-quarantine may be voluntarily observed in the home environment. Therein, administrative orders always have priority, but can and should be supplemented by voluntary and employer-initiated measures.

## Material and methods

### Organizational implementation

The SARS-CoV2 isolation and quarantine management at Marl Chemical Park is directed centrally by the plant medical service of Evonik Industries AG. It applies to a large number of companies and sub-contractors and is coordinated with the responsible municipal health departments and authorities. A digital process management called “corona control center” is established for this aim. Results in this paper are developed and tested in this context.

A central part of these results is an Excel dashboard that facilitates risk assessment and strategy planning. This has been implemented in Excel, as it is a convenient tool for most occupational health departments and requires no further installation.

### Epidemiological formulas

The formulas for isolation and quarantine management therein rely on and are assessed using the fundamental research of Lipschik et al. 2003 [1] and Ferretti et al. 2020 [2] and are evolved in the following:

The basic reproduction factor *R*_0_ describes how many persons on infected person infects in average. According to Lipschik et al. 2003 [1] one can calculate it via the formula

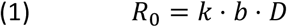

where *k* is the average number of single contacts per day, *b* is the infection probability per contact and day and *D* ist the duration of infectiousness (in days).

The reproduction factor influenced by the intervention (*R*_*int*_) is an indicator of success for the intervention [1]:

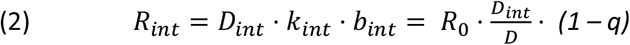

Thus, the success of a protective measure depends on the reduction of the number of infectious contact days by the intervention (*D*_*int*_: remaining number of infectious days with contacts under the intervention) and the infection probability reduction *q* (where 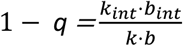; via additional protec-tive measures, testing strategies or general contact restrictions).

These formulas are valid for diseases with a constant infectivity during the course of the disease, whereas considerable differences of infectivity are present during the course of a SARS-CoV-2 infection. An approximation of the infection probability on each single day can be calculated using the epidemiologic models in Ferrati et al. 2020 [2]: therein, generation time (i.e. the duration between one infection and a subsequent infection) is shown to follow a Weibull distribution with shape parameter *k* = 2.886 and scale parameter 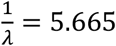 which yields the following density function *β*(*τ*) (for *τ* > 0, where *τ* = 0 is the time of infection) for mean infectivity over time:

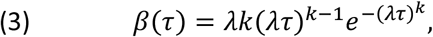

Cumulated infection probability in the time interval [*t*_1_,*t*_2_] (in days since the infection of the source case) thus is

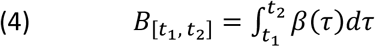

In this formula, the day-specific infection probabilities are calculated based on the time of the infection of the source case (t=0). In practice, calculations referring to the time of infection are hard to implement, as in general it is not known. This is why it is more convenient to use the symptom onset, which is day 5 after the infection according to [2], as a point of reference for the calculation of day-specific risks. In some cases, the information on the date of symptom onset is not available (or the case is asymptomatic) and thus needs to be replaced by the day of the positive testing (which is assumed to be on day 1 after symptom onset).

Consequently, formula (4) needs to be adapted to

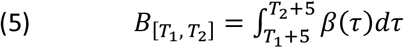

where *T* = *t* − 5 represents the time since symptom onset in days.

Here, the events “infection time” and “symptom onset” are standardized to noon in the calculations and the time-specific infection probabilities are cumulated to weekdays (Fig. 1).

**Fig. 1:**
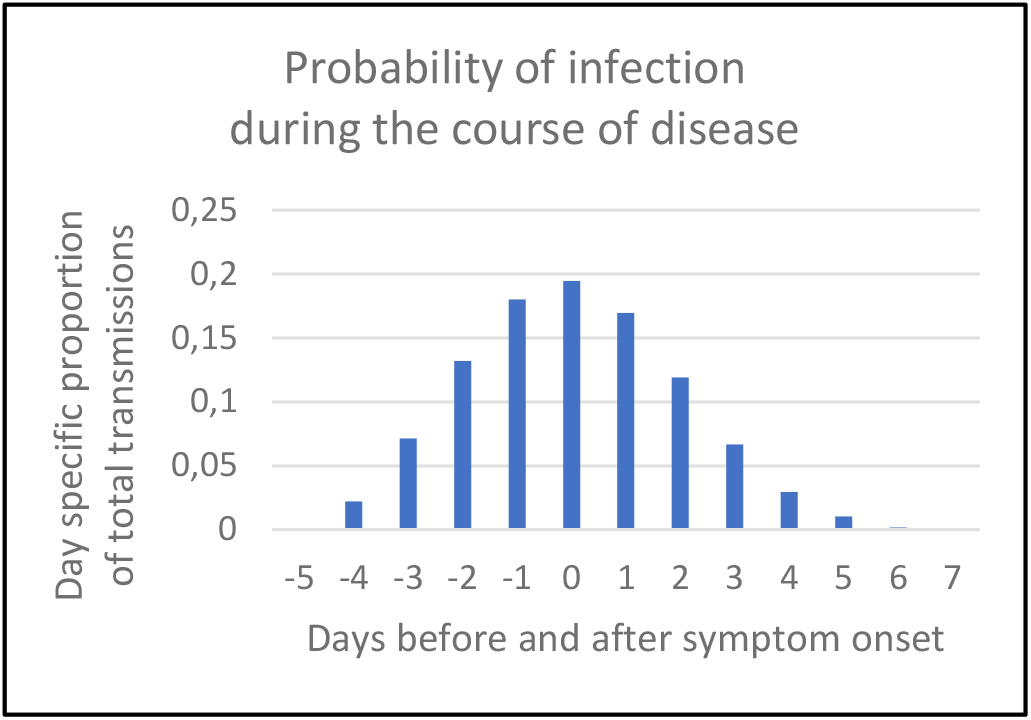
Day-specific proportion of total transmissions on different (working) days, as referring to symptom onset (day 0)

Formula (5) thus simplifies to

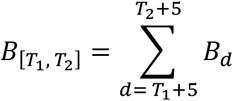

Where

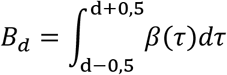

is the cumulated infection risk on day d.

Here, the question arises whether this distribution oriented at symptom onset is influenced by the incubation time. He et al. 2020 [3] showed that the probability of infection transmission is very low until day −4 and experiences a steep increase on day −3, independently of incubation time. Thus, the calculation of daily infection probabilities is valid as a good approximation of the individual incubation time according to the current literature.

The effectivity *E*_*int*_ of the respective isolation or quarantine measures taken on day *i* can be calculated from formula (4) as

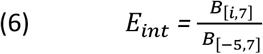

i: day after the implementation of the quarantine or isolation measure

The reproduction number taking account of the respective levels of intervention can be calculated by the following adaption of formula (2):

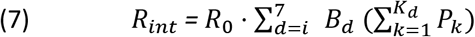

*R*_int:_ modified reproduction number after the intervention, *R*_0_: basic reproduction, *K*_*d*_: number of contacts on day d, *P*_*k*_: transmission probability of to the respective contact, *B*_*d*_ = *B*_[*d*−0,5;*d*+0,5]_ : cumulated infectivity on day d.

Formula (7) is a more detailed variant of formula (2), as it takes the contact day specific infectivity *B*_*d*_ into account instead of the number of infective days 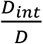 only. Instead of the cumulated risk reduction (1 − *q*) in formula (2), formula (7) works with the number of contacts *K*_*d*_ on each single day, which is reduced by an intervention, as well as the contact specific infection probability *P*_*k*_ (which can depend on vaccination status, mask use, ventilation and distance between the two persons).

### Individual risk assessment

Social contacts are identified by interviewing the employees, and in some cases by an additional inspection of contact diaries. In individual cases, digital media such as tracking systems could be considered as a supplementary source of information. The assessment of individual infection risks to three categories (high, medium and low risk) was implemented using simple schemata that were designed according to the general recommendations of the health authorities for estimating the main factors contact distance and contact time. For plant specific transmissions, risk levels for high, medium and low risks were assessed as 0.2, 0.1 and 0.05 respectively. These values must be adapted to different transmission conditions (which are influenced by factors such as room size, ventilation conditions, percentage of indoor work, type of work, etc.).

The basic reproduction number *R*_0_ shows a high level of uncertainty in different studies [4,5,6,7]. For the calculations herein we use *R*_0_ = 2.5 for the wild type of Covid-19, which corresponds to the best available estimate according to the Pandemic Planning Scenarios of CDC [8]. Concerning virus variants, the value of *R*_0_ is adapted to recent estimates of the respective infectivity [9,10] (B 1.1.7. *R*_0_ =3,75). The expected declines in the baseline reproduction number due to increasing immunization of the population must be accounted for in the future by adjusting the values of *R*_0_.

### Diagnostic tests

For diagnostic tests, the SARS-CoV-2 Rapid Antigen Test by SD Biosensor, Korea, distributed by Roche Diagnostics GmbH was used. Its sensitivity is 96,52 % (95 % CI 91,33 – 99,04 %) and its specifity 99,68 % (95 % CI 98,22 – 99,99 %). Samples were obtained with deep nasopharyngeal swabs.

## Layout in practice and results

Comparisons of single strategies of outbreak containment such as isolation, contact tracing with quarantine, symptom monitoring and testing strategies in the literature show that combinations of different measures can be considerably more effective than single measures [11, 12]. The excel based quarantine calculator named “ChainCUT” has been developed in order to be able to manage the increased complexity of this success-optimized approach.

After recording the identified contacts, the effectiveness of the interventions is calculated and infection risks are analyzed for the specific operational situation and virus variant. Based on this risk calculation, combined measures for the containment of intra company infection chains are calculated.

Risk assessments and extended measures are therefore carried out not only for the source case as virus generation 0 and the direct contacts as generation 1, but also for the indirect contacts (generation 2), i.e. all persons having had no contact to the source case, but contact to its direct contact persons. By extending the observation and interventions, further spread (generation 3) can be prevented even for virus variants with increased infectivity (Fig. 2).

**Fig. 2:**
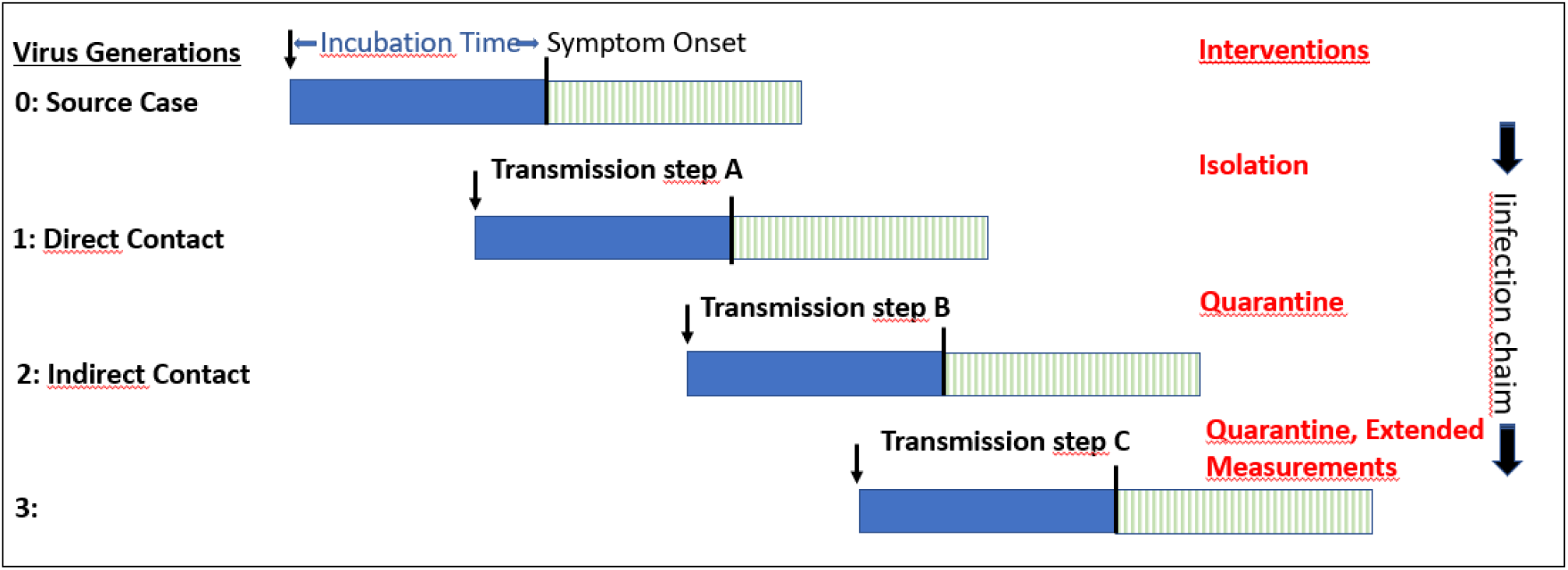
Overview of the infection chain with virus generations, transmissions and interventions.

In order to capture as many infection risks as possible, contact detection will be extended to day −3, a day with medium-high infectivity (Fig. 1). If the onset of symptoms is unknown, the determination of day 0 is uncertain. Here, the inclusion of day −3 can minimize the impact of possible classification errors.

Regarding common stays in the work or social area, the risk of infection cannot be excluded even if the minimum distance is respected. Social area is defined as a simultaneous stay in a common area such as break rooms, smoking rooms, washrooms or changing rooms. In this context, low risks due to stays in the same indoor rooms, occasional brief encounters, accidental exposures due to air currents or smear infections are assumed and taken into account on a rough basis.

The ChainCUT quarantine calculator visualizes the effectivity of the separation measure (using calculations based on formula 6) and the reproduction numbers after the intervention (according to formula 7) in a risk analysis dashboard separately for the different intervention levels (cf. fig. 3 and 4 in the appendix), in order to allow for a precise control of the measures.

**Fig. 3 a:**
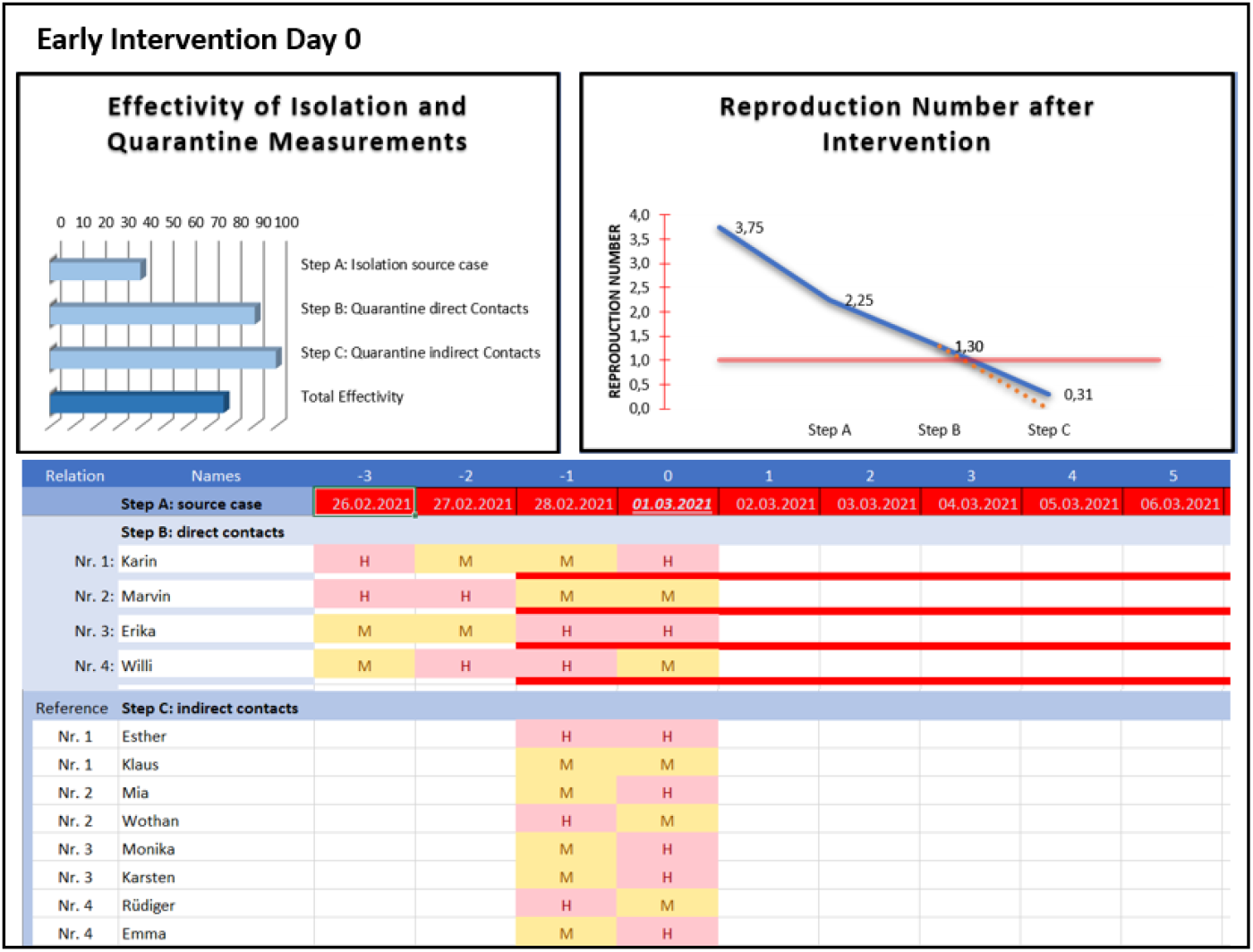
Initiation of the source case’s isolation and the quarantine of the contact persons directly after symptom onset. *E*_*int*_ (top left) calculated from formula 6. *R*_*int*_(top right) calculated from formula 7. The dashed line indicates the R value after the negative testing of all direct contact persons. The contact table contains high (H) and medium (M) contagion risk contacts. Above are the contacts between the source case and its direct contacts, below the contacts of the direct and indirect contacts.

**Fig. 3 b:**
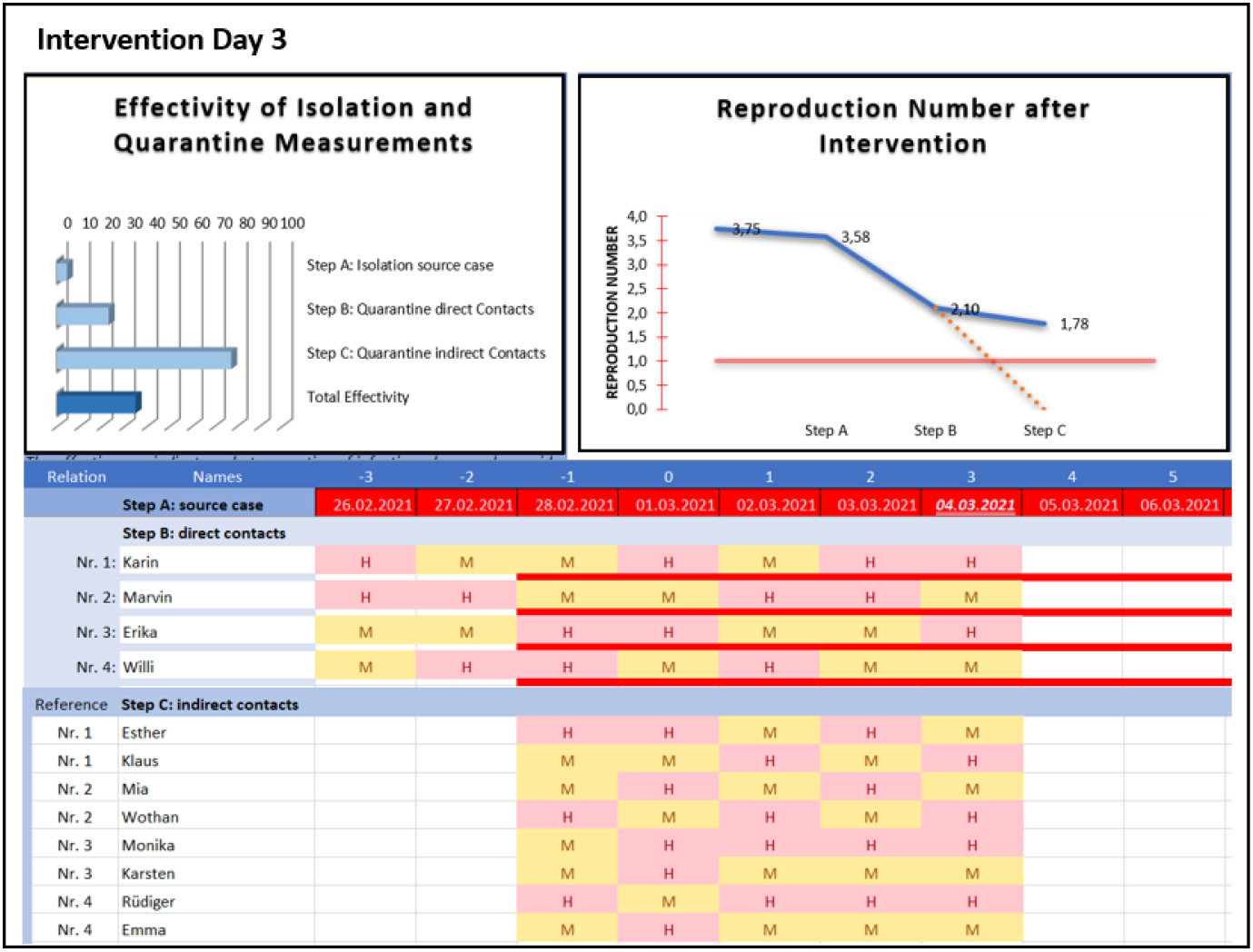
Initiation of the source case’s isolation and the quarantine of the contact persons on day 3 after symptom onset. Contacts are continued until day 3.

**Fig. 3 c:**
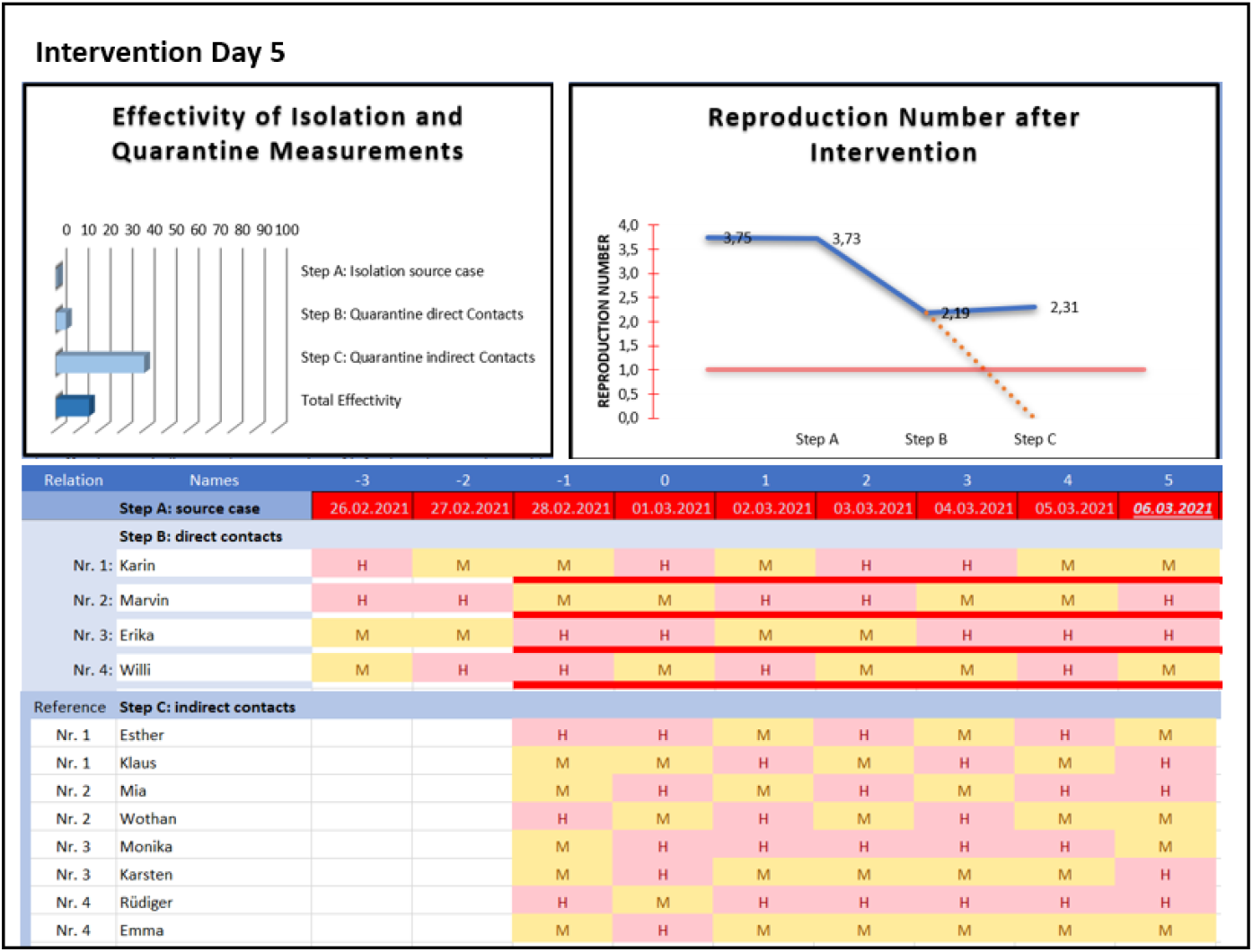
Initiation of the source case’s isolation and the quarantine of the contact persons on day 5 after symptom onset. Contacts are continued until day 5.

**Fig. 4:**
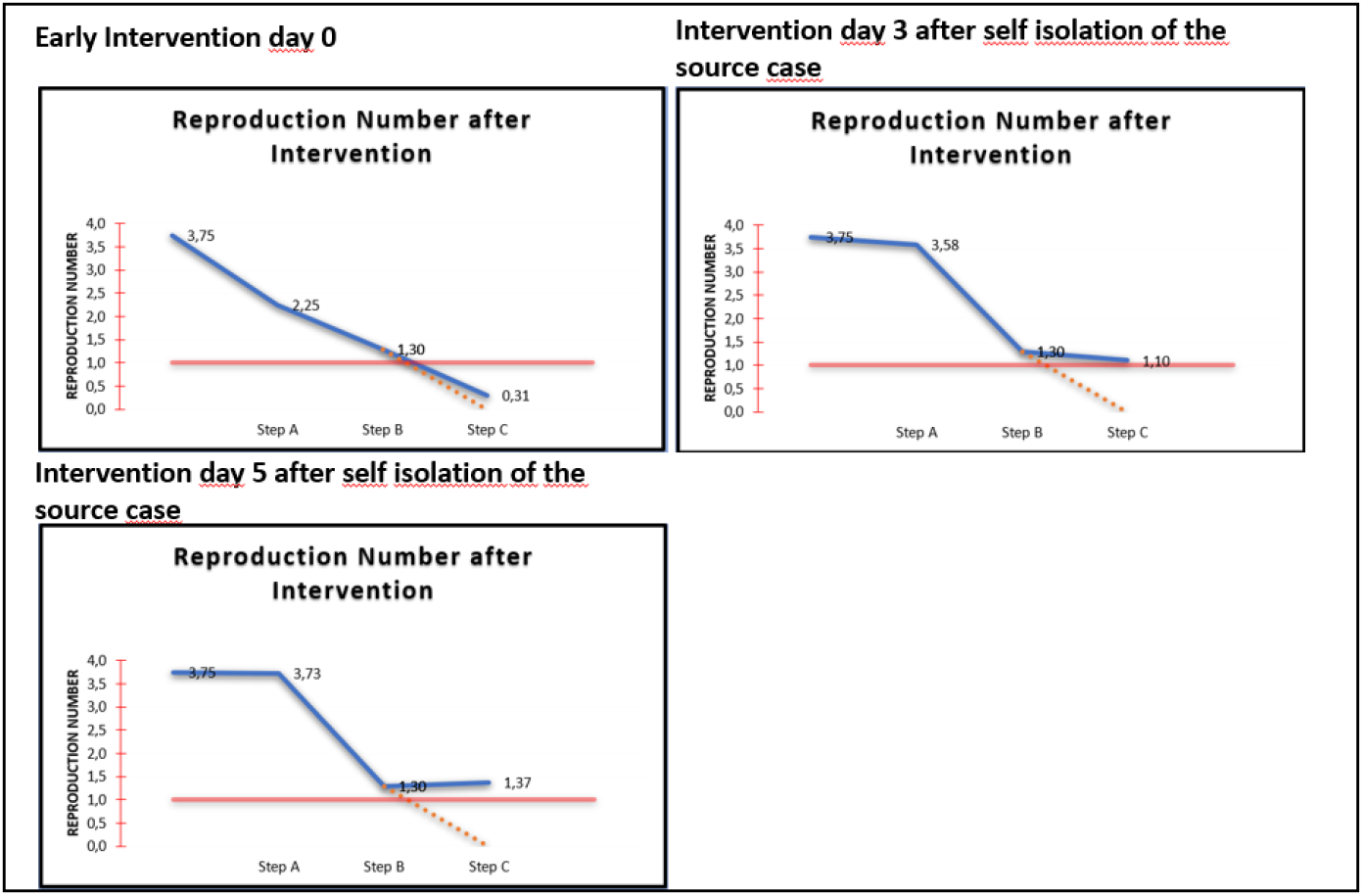
Comparison of reproduction values in early intervention on day 0 and in interventions on day 3 and 5, if a self-isolation of the source case on day 0 without subsequent contacts of the source case to the direct contact persons and with continued contacts of the direct contact persons to the indirect contact persons is present.

In example calculations, the minimizations of the time lag in initiating measures could be identified as a determining success factor for isolation and quarantine management. One of these example calculations assumes that the source case has four direct contact persons per day, with a total transmission risk of 0.6 (2 contacts with high infection risks, 2 contacts with low infection risk). For each of these contact persons, two indirect contacts with an infection risk of 0.3 are assumed (cf. fig. 3 and 4). This calculation is performed for the implementation of preventive measures on day 0 (fig. 3a), day 3 (fig. 3b) and day 5 (fig. 3c) respectively. Day 0 is the day the first symptoms occur. Time delay until day 3 is the typical case, where the result of PCR tests are waited for before the implementation of the measures. If additional delays occur prior to the intervention, frequently day 5 is reached.

In the case of the early intervention on day 0, the example calculation for B 1.1.7 with basic reproduction number 3.75 shows a reproduction factor of 2.25 for the source case (level A) after the isolation measure, for the direct contacts (level B), a reproduction factor of 1.30 after quarantine, and for the indirect contacts (level C), of 0.31 after quarantine. Thus, despite the extensive contacts, the R-factor in level C is clearly below 1 and the risk of an infection chain to occur is low. With a time delay resulting in an intervention measure start on day 3, the R factor for level C is 1.78 and thus shows the risk of a continued infection chain (cf. fig. 3c). If the intervention starts on day 5, the R factor for level C is 2.31. Risk containment to a small group of persons is hardly possible in the latter case.

Assuming self-isolation of the source case on day 0 without any further contacts, the time delay concerning its direct in indirect contact persons results in an R value of 1.10 (assuming full intervention start on day 3) or 1.37 (assuming full intervention start on day 5) (cf. fig.4 in the appendix). This implies that every delay in the implementation of the measures reduces considerably the chance of an early containment of the infection incident; this is particularly true for virus variants with increased infectivity.

The validity of the calculated reproduction values can be significantly improved by combining them with a differentiated and targeted testing strategy as an extended measure (app. fig. 3 dotted line). In this setting, the aim is to perform a rapid antigen test for direct contacts on the day the measures are initiated. This can be used to determine whether direct contacts are infectious at the start of their quarantine with a high degree of certainty. In the case of negative test results, an infection transmission to the respective indirect contact persons can be excluded. If all test results are negative (R value in level C: 0.00), the containment of the infection chain can be concluded with a high level of certainty. In this case, no further quarantine measures for indirect contacts need to be initiated. The negative result of the rapid antigen test for direct contact persons excludes their infectivity, but the aim of the testing is explicitly not to exclude an infection and thus to avoid quarantine for direct contacts.

In the case of uncertain information in the contact questioning, or a high number of persons without direct contact, but within the same work and social area, an increased infection risk for the whole group is possible, especially concerning virus variants with increased infection risks. This is why a temporary regular rapid antigen testing of the whole group can be required in this case.

## Limitations and uncertainties

The calculation of the infection probabilities is oriented to symptom onset, but in practice, the day of symptom onset is not always known, due to a lack communication of these data by the person concerned or symptomless testing for various reasons, for instance. The determination of day 0 is thus subject to uncertainties. A misidentification can result in an incomplete consideration of the risks on highly infective days (day −2 to +1), and thereby to a decrease in effectivity of the measures. The systematic consideration of day −3 can minimize, but not eliminate, this effect.

In the contact tracing by interviews, the motivation and recall capacities of the employees are a key factor. If the measures are instantly implemented, recall capacities are a minor factor, but the motivation to communicate a contact event can be impaired if ordered protective measures were not complied to by the employee and there is a lack of a sufficient culture of trust within the company from the point of view of the employee.

The estimate of the virus intake of the receiver according to the transmission conditions and the effect of protective measures is key to the calculation of transmission risks and can be complex, especially in a work environment. In general, simple estimation schemes by the health authorities are used, which essentially take the distance and the duration of a contact into account. Observations in practice show that in general, distances are overestimated and contact durations are underestimated. The cumulative consideration of different contacts, transmission conditions and indoor exposures for the total work duration is a special challenge. In total, frequent misinterpretations of infection risks have to be expected and accounted for according to practical experience. The development and implementation of practicable transmission simulation tools can lead to significant improvements of the risk estimates.

## Discussion

The secure prevention of intra company infection chains, with special regards to virus variants with increased infectivity, requires a complete risk determination and assessment. The qualified risk analysis identifies optimal measures to contain outbreaks at an early stage and to keep the number of persons at risk small. The presented epidemiologic modelling calculates infection risks and the expected success of the measures across virus generations. The informative value of these calculations can be significantly enhanced by the combination with a differentiated testing strategy.

The registration of contacts with an infectious source at the workplace is the basis of the calculation of cumulated transmission risks for each pair of source and receiver. In classic contact tracing, only persons having a high risk contact (defined by the unprotected stay within the minimal social distance for a predefined duration) are eligible for quarantine measures. Therein, risks caused by multiple contacts on different days below the definition of a high risk contact are systematically not taken into account. This leads to gaps in the risk assessment and, as follows, in the selection of measures. The presented model evaluates cumulatively all contacts depending on the respective contact-specific transmission risk and the day-specific infectivity of the source. In the calculations, contacts on day −3 with a medium-high infectivity are additionally considered. Thus, a complete picture of the risks for each source-receiver pair is attained.

For infectious diseases with a high contagion risk before symptom onset, epidemiologic publications [13, 14] describe the great importance of the time lag until measures for outbreak containment are initiated. The results of the example calculation according to fig. 3 and 4 confirm this relationship. At the onset of the first symptoms of the source case on day 0, depending on the number of contacts and their probability of transmission, transmission from virus generation 0 (source) to generation 1 (direct contacts) must already be expected (fig. 2). A further transmission to generation 2 (indirect contacts of the source) is still improbable at this point. The intervention measures at this point are therefore promising and show reproduction values far below 1 in the example calculation for indirect contacts for B 1.1.7.

Every additional day until the implementation of the interventions increases the probability of transmission to generation 2 (indirect contacts): The example calculation shows the R-value 1,78 (assuming intervention start on day 3) and even 2,31 (assuming intervention start on day 5) respectively, for B 1.1.7. This considerably expands the group of persons at infection risk, as well as the scope of necessary measures.

In general, health authorities are informed via positive PCR testing and take action thereafter, which is why a delay of three days after symptom onset has to be considered as the ideal case in practice. In general, the initiated interventions refer to the source case and its direct contact persons. As, at this point in time, a transmission to the next generation has to be expected, a fast containment of the infection chain cannot be assumed. This is especially true for virus variants with increased infectivity.

For this reason, the earliest possible implementation of measures by the company is a decisive success factor, especially with regard to the new virus variants. The timely execution of rapid antigen tests in case of any suspicion has to be initiated by the company. Waiting for testing and results by the health system often leads to significant time delays in practice.

The use of intra-company tracking systems or contact protocols can facilitate the identification of contacts. However, the use of such a system alone does not constitute an efficient risk management, as the factors “reaction time until the implementation of measures” and “consideration of all risks” have a predominant impact on the success of the measures.

Extended measures, such as a differentiated and targeted testing strategy for the direct contact persons at the beginning of the quarantine are necessary when an elevated reproduction value in level C is present. With a rapid antigen testing of contact persons, in case of a negative result, the infection transmission to the respective indirect contact persons can be excluded with a high level of certainty. In the case of a positive result, quarantine measures can be limited to the indirect contacts concerned. Using these extended measures, the number of persons at risk can be limited, preserving personnel resources and avoiding mass testing and quarantines. The testing can be extended to the whole work group for a predefined period of time in case of unclear intra-group contact constellations with many persons within the same work and social area or unclear statements on contacts. In principle, it is advantageous to perform targeted testing for persons and groups with elevated infection risks, due to the better information content and prediction value.

When the operational staff availability critically decreases due to quarantine measures, alternative measures can be implemented. An alternative to quarantine is the organization of single work or isolated work. This requires the spatial and organizational separation of the work place of the respective person avoiding any social contacts. If this is not possible a daily testing prior to the work day can be used as a substitute for quarantine. In this case, special requirements to the quality and information content of the rapid test must be set, as well as to the professionally performed sampling. Due to the prevailing residual risks, complete compliance to the basic protective measures is indispensable.

The ChainCUT quarantine calculator performs an automatic risk analysis after the recording of all risk contacts and suggests individual measures. This provides workplace managers with an easy-to-use tool offering the medical and epidemiological expertise needed to make decisions and to initiate optimal interventions. In combination with the organization of a rapid testing initiated by the company, all time critical measures can be initiated as early as possible. Thus, this tool is systematically outperforming expert-based time-delayed approaches for the rapid containment of outbreaks.

## Conclusion

Due to their high infectivity, the novel virus variants of SARS-CoV-2 can cause significant outbreaks especially at the workplace, putting employees as well as the continuation of business activities in danger. In this context, facilities and employers have very effective intervention options at their disposal, such as the organization and binding directive of protective measures, as well as the segregation of employees by refraining from on-site work performance. Using these intervention possibilities and an evolved risk management discussed in this paper, it is possible to minimize intra-company risks of contagion despite the social contacts associated with business activities.

The targeted risk-based combination of basic protective measures, workplace isolation and quarantine management, symptom monitoring and a testing strategy shows a significantly higher effectivity than single measures. This efficiency has to be optimized using a by a forward directed separation strategy over several virus generations and early interventions, especially in the context of critical virus mutations. An epidemiologic model allowing for a differentiated risk analysis for contact persons based on the day-dependent infectivity is the rationale of these measures. Thus, risk assessment can be improved and targeted measures can be derived.

Example calculations show that the timing of the start of interventions is a crucial factor of success. In practice, contact tracing by the authorities can often only start with a significant delay, which is why immediate workplace interventions are a decisive improvement in the rapid containment of outbreaks. That quarantine calculator derived from these principles enables even non-specialists to perform a differentiated risk analysis and to introduce optimized measures. Targeted testing routines and alternative measures ensure staff availability.

## Software

The “ChainCUT quarantine calculator” and the “aerosol indoor simulator” are available as Excel applications on www.evonik.com/corona for free use.

## Data Availability

all data and sources are listed in the references of the manuscripts

## Appendix

## References

1. Lipsitch, M., Cohen, T., Cooper, B., Robins, J.M., Ma, S., James, L., Gopalakrishna, G., Chew, S. K., Tan, C.C., Samore, M.H., Fisman, D. Murray, M. (2003). Transmission Dynamics and Control of Severe Acute Respiratory Syndrome. Science. 2003 June 20; 300(5627): 1966–1970. DOI:10.1126/science.1086616.

2. Ferretti, L., Wymant, C., Kendall, M., Zhao, L., Nurtay, A., Abeler-Dörner, L., Parker, M., Bonsall, D., Fraser, C. (2020). Quantifying SARS-CoV-2 transmission suggests epidemic control with digital contact tracing. Science 10.1126/science.abb6936. DOI: 10.1126/science.abb6936.

3. He, X., Lau, E.H.Y., Wu, P. Deng, X., Wang, J., Hao, X., Lau, Y.C., Wong, J.Y., Guan, Y., Tan, X., Mo, X, Chen, X., Liao, B., Chen, W., Hu, F., Zhang, Q., Zhong, M., Wu, Y., Zhao, L., Zhang, F., Cowling, B.J., Li, F., Leung, G.M. (2020). Temporal dynamics in viral shedding and transmissibility of COVID-19. Nature Medicine, VOL 26, 672 MAY 2020, 672–675. DOI: 10.1038/s41591-020-0869-5.

4. Sanche, S., Lin, Y.T., Xu, C., Romero-Severson, E., Hengartner, N., Ke, R. (2020). Contagiousness and Rapid Spread of Severe Acute Respiratory Syndrome Coronavirus 2. Emerging Infectious Diseases, www.cdc.gov/eid, Vol. 26, No. 7, July 2020. DOI: 10.3201/eid2607.200282.

5. Imai, N. Cori, A., Dorigatti, I., Baguelin, M., Donnelly, C.A., Riley, S., Ferguso, N.M. (2020). Report 3: transmissibility of 2019-nCov. Imperial College London (25-01-2020), DOI: 10.25561/77148.

6. Li, Q., Guan, X., Wu, P., et al. (2020). Early transmission dynamics in Wuhan, China, of novel coronavirus-infected pneumonia. N Engl J Med. 2020;382(13):1199–1207.

7. Kucharski, A.J., Russell, T.W., Diamond, C., et al.: Early dynamics of transmission and control of COVID-19: a mathematical modelling study. medRxiv 2020

8. CDC, Pandemic Planning Scenarios 10.03.2021, www.cdc.gov/coronavirus/2019-ncov/hcp/planning-scenarios.html

9. Vöhringer, H., Sinnott, M., Amato, R., Martincorena, I., Kwiatkowski, D., Barrett, J.C., and Gerstung, M. (2020). Lineage-specific growth of SARS-CoV-2 B.1.1.7 during the English national lockdown.

10. Volz, E., Mishra, S., Chand, M., Barrett, J.C., Johnson, R., Geidelberg, L., Hinsley, W.R., Laydon, D.J., Dabrera, G., O’Toole, á., et al. (2021). Transmission of SARS-CoV-2 Lineage B.1.1.7 in England: Insights from linking epidemiological and genetic data. medRxiv, 2020.2012.2030.20249034.

11. Kucharski, A.J., Klepac, P., Conlan, A.J.K., Kissler, S.M., Tang, M.L., Fry, H., Gog, J.R., Edmunds, W.J. (2020). Effectiveness of isolation, testing, contact tracing, and physical distancing on reducing transmission of SARS-CoV-2 in different settings: a mathematical modelling study. Lancet Infect Dis. 2020 Oct;20(10):1151–1160. DOI: 10.1016/S1473-3099(20)30457-6. Epub 2020 Jun 16.

12. Peak, C.M. Kahn, R., Grad, Y.H., Childs, L.M., Li, R., Lipsitch, M. Buckee, C.O. (2020). Comparative Impact of Individual Quarantine vs. Active Monitoring of Contacts for the Mitigation of COVID-19: a modelling study. DOI: 10.1101/2020.03.05.20031088.

13. Fraser, C., Riley, S., Anderson, R.M., Ferguson, M. (2004). Factors that make an infectious disease outbreak controllable. PNAS, April 20, 2004, vol. 101, no. 16, 6147. https://www.pnas.org/content/101/16/6146.short

14. Hellewell, J., Abbott, S., Gimma, A., Bosse, N.I., Jarvis, C.I., Russell, T.W., Munday, J.D., Kucharski, A.J., Edmunds, W.J., Funk, S., Eggo, R.M. (2020). Feasibility of controlling COVID-19 outbreaks by isolation of cases and contacts. www.thelancet.com/lancetgh Vol 8 April 2020.

